# Characterization of motor function recovery using muscle synergies in stroke patients undergoing upper limb rehabilitation

**DOI:** 10.1101/2024.09.24.24314287

**Authors:** Giorgia Pregnolato, Giacomo Severini, Lorenza Maistrello, Daniele Rimini, Tiziana Lencioni, Ilaria Carpinella, Maurizio Ferrarin, Johanna Jonsdottir, Vincent C. K. Cheung, Andrea Turolla

## Abstract

In stroke rehabilitation, muscle synergies have been investigated to provide information on the level of upper limb motor impairment, but not yet for explaining motor recovery after therapy. In this study, we analysed muscle synergies parameters extracted from 62 people with stroke who underwent a specific upper limb treatment (20 sessions, 1h/day, 5d/week, 4 weeks) consisting of virtual reality, robotic or conventional treatment. Overall, participants improved upper limb motor function (Fugl-Meyer Assessment Upper Extremity-FMA-UE: Δ= 7.14 ± 7.46, p< 0.001) but the number of muscle synergies of the impaired side (N-aff) did not change after treatment (N-aff: T0= 8.8 ± 1.5; T1=8.7 ± 1.4; p=0.374). Then, we stratified the sample into Responder (No.=34) and Non-Responder (No.=28) participants, based on the Minimal Clinical Important Difference value of FMA-UE (Δ >5 points). We investigated merging and fractionation indexes in both subgroups and only the Responder subgroup significantly decreased the percentage of merged muscle synergies (p=0.004). No significant changes in the fractionation index resulted in either subgroup.

Finally, we identified vocabularies of affected upper limb motor synergies, before (No. 13 synergies) and after treatment (No. 14 synergies), and in unaffected upper limb (No. 16 synergies). We identified upper limb motor functions associated with each muscle synergy in each vocabulary based on the muscles represented in the muscle synergy. There were no differences in motor functions characterizing Responder patients. However, after therapy, both Responder and Non-Responder subgroups were characterized by the same distribution of motor functions across muscle synergies observed in the unaffected limb.

**Trial registration:** The trial is registered in ClinicalTrial.gov, identifier number NCT03530358 (https://clinicaltrials.gov/study/NCT03530358).

**Key messages:**

- Rehabilitation therapy for the upper limb induces reduction of muscle synergies merging in people with stroke expressing clinically important improvement of motor function. This muscular pattern is in accordance with motor control mechanisms underpinning functional recovery.
- Motor function of the affected upper limb at baseline did not characterize the muscular profile of patients responding to rehabilitation treatment (Responder).
- After therapy, all patients regardless the amount of motor function improvement (i.e., Responder, Non-Responder) express a muscular profile of the affected upper limb like the unaffected upper limb.

## Introduction

Impairment of upper limb (UL) motor function is one of the most common sequelae after stroke (1), with 50% of survivors still experiencing disabling consequences at 6 months from onset (2, 3). In clinical practice, the use of valid assessment tools of UL motor function is recommended to improve the prognosis of recovery and to measure the effectiveness of personalized-specific rehabilitation treatments (4–7). A large number of clinical outcome measures are used to assess the UL motor function (5, 6). Moreover, the use of instrumental measurement methods (e.g., kinematic, electromyography) in clinical settings is strongly recommended (8, 9).

The muscle synergy model has been proposed to extract quantitative indexes associated with neurophysiological patterns of voluntary motor execution (10, 11). In the muscle synergies model the control of voluntary movements is postulated to be generated by the linear combination of pools of muscles organised into modules (i.e., synergies) (11). Muscle synergy features (e.g., time-invariant modules and their relative activations weights) are usually extracted applying decomposition algorithms to the surface electromyography (sEMG) signals recorded during execution of voluntary motor tasks (10, 12). A single synergy is responsible for activating a group of muscles each with a specific weight. Voluntary movements result from the linear combination of synergies, with varying intensity and timing (12).

In stroke rehabilitation, muscle synergies have been investigated as a means to characterize the level of UL motor impairment, their coherence with clinical outcomes and with kinematic features (13–19). Evidence suggests that muscle synergies reflect the activity of spinal interneurons, and recent studies outlined that after stroke muscles synergies parameters are consistent across individuals regardless the UL (i.e., affected, unaffected) they are extracted from (13, 20). Thus, the impairment of motor function following stroke could be described by variability in the modules activation (i.e., weight of each muscle contribution as a module unit) (21).

Cheung et al. distinguished three patterns of UL muscle synergies modules reorganisation after stroke, which appear to correlate with the clinical characteristics of survivors (13). The first pattern is related to synergy preservation, which occurs when then same module is extracted from both the unaffected and stroke-affected UL (13). The second pattern is the synergy merging, which occurs when a synergy from the stroke-affected UL can be described as a combination of two or more modules extracted from the unaffected UL (13, 21). The merging rate was reported to increase in accordance with the severity of UL motor function impairment and significantly different between severe and mild impairments (22). The third pattern is the synergy fractionation, which occurs when two or more synergies extracted from the stroke-affected UL can be combined to constitute a synergy extracted from the unaffected UL (13, 21). The rate of muscle synergies fractionation after stroke positively correlates with time from onset, suggesting that for execution of voluntary movements in the chronic phase of recovery new UL muscle synergies are developed, that are simplified versions of unimpaired synergies (13).

Despite relationships between clinical features and synergy patterns (i.e., preservation, merging and fractionation) were explored in the literature, evidence is still low with regard to changes of muscle synergy features and patterns following specific UL training, after stroke.

In this study, we investigated the effect of a specific UL rehabilitation programme consisting of 20 sessions of conventional, robotic-assisted or virtual reality-based training, in people with stroke (PwS). We analysed clinical outcome measures and muscle synergies features extracted before and after UL training, also comparing muscle synergies features between participants responding (i.e., Responder) and not responding (i.e., Non-Responder) to UL therapy, as defined by clinical outcome measures.

## Methods

The study is a secondary analysis of the trial “Modularity for Sensory Motor Control” (MO-SE), whose protocol was registered in ClinicalTrial.gov (ID NCT03530358).

### Participants

Participants enrolled were inpatients at the San Camillo IRCCS Hospital (Venice, Italy) and the IRCCS Santa Maria Nascente at Don Carlo Gnocchi Foundation (Milan, Italy). All participants were informed on the aims and modalities of the study and provided an informed written consent. Inclusion criteria were adult participants (older than 18 years), diagnosis of unilateral stroke (haemorrhagic or ischemic), impairment of UL motor function defined as a score between 5 and 61 points at the Fugl-Meyer Assessment scale for Upper Extremity (FMA-UE). Moreover, patients were excluded in case of severe cognitive impairment, severe impairment of verbal comprehension or untreated seizures.

### Study design

This secondary analysis of the study was designed with a prospective, longitudinal, and interventional approach. Participants underwent a rehabilitation treatment targeting the UL delivered 1 hour a day, 5 times a week, for 4 weeks (20 sessions). In each medical centre involved, after enrolment, participants were allocated randomly to an experimental, or conventional treatment (CT). At the San Camillo IRCCS Hospital (Venice, Italy) the experimental modality was based on virtual reality (Virtual Reality treatment, VRT) and delivered by the Virtual Reality Rehabilitation System (VRRS®, Khymeia Group Ltd., Padova, Italy). In the VRT group patients performed diverse reaching exercises by interacting with a virtual environment using a 3D motion-tracking system (Polhemus 3Space FasTrack, Polhemus, Colchester, VT, USA) with a sampling frequency of 120 Hz. The sensor was fixed on the hand back of the impaired UL by a glove worn by the patient. At the IRCCS Santa Maria Nascente at Don Carlo Gnocchi Foundation (Milan, Italy) the experimental modality was robotic-assisted (robotic treatment, RT) and provided by the Braccio di Ferro (Celin s.r.l., Follo, SP, Italy) device, an haptic manipulandum designed to assist or perturb centre-out biplanar reaching movements, initiated and actuated by the patients (23). During the treatment, the participant instructed to perform planar movements on a table to reach specific targets displayed on a screen. In both centres, the conventional treatment (CT) consisted of an UL programme based on passive mobilization, active movements, and task-oriented exercises.

### Outcome measures

To detect the effect of UL rehabilitation, patients were assessed before and after each treatment by a set of outcome measures administered by physiotherapists with expertise in neurological physiotherapy.

The primary outcome of the study was the FMA-UE, a reference standard for the assessment of UL gross motor function after stroke (24, 25). The FMA-UE is composed by 33 items each one scored as follow: 0 point = the patients cannot perform the required task; 1 point = the patient can perform the task but incompletely; 2 points = the patient can perform the task. Overall, total FMA-UE score ranges from a minimum of 0 (no motor function) to a maximum of 66 (normal function) points.

Moreover, secondary outcome measures were considered. The FMA, sensation (FMA-S) was used to quantify the level of sensorimotor impairment in the affected UL. The FMA-S is composed by 12 items with the following scoring system: 0 = no sensation, 1 = incorrect sensation, 2 = preserved sensation. Overall, the FMA-S score ranges from a minimum of 0 (no sensation) to a maximum of 24 (normal sensation) points. The FMA, pain and range of motion (FMA-P) was used to assess the pain and the passive range of motion of joints in the affected UL. The pain scoring is: 0 = severe pain, 1 = moderate pain, 2 = absence of pain. The scoring for the passive range of motion is: 0 = severe limitation, 1 = moderate limitation, 2 = normal. Overall, the FMA-P score ranges from a minimum of 0 (no impairment) to a maximum of 48 (normal functions) points. The Modified Ashworth Scale (MAS) (26) was used to assess the muscle tone response to manual stretch reflex. The MAS was tested in five key muscles of the affected side (i.e., pectoralis major; biceps brachialis; flexor carpii; flexor digitorum profundus; flexor digitorum superficialis). MAS is an ordinal scale of 6 ranks (0; 1; 1+; 2; 3; 4), with 0 representing normal muscle tone, and 4 severe muscle spasticity. The MAS maximum score is 20 points.

The Functional Independence Measure (FIM) (27) was used to assess the level of the independence in daily activities. The FIM is composed by 18 items, grouped in 6 sections: self-care; sphincter control; transfers; locomotion; communication; social and cognition. Each item can be scored from 1 to 7 (1 = total assistance; 2 = maximal assistance; 3 = moderate assistance; 4 = minimal assistance; 5 = supervision; 6 = modified independence; 7 = complete independence). The FIM score ranges from a minimum of 18 points (full dependence) to a maximum of 126 points (full independence).

Demographic and anamnestic data included: age, sex, type of lesion (i.e., ischaemic, or haemorrhagic), lesioned hemisphere (i.e., left or right) and time from stroke onset.

### sEMG acquisition

The sEMG signal was acquired from 16 UL muscles, following the SENIAM guidelines for sensors location (28). The muscles sampled were: the triceps brachii, medial (TME) and lateral (TLA) heads; the biceps brachii, short (BIS) and long (BIL) heads; the deltoideus anterior (DEA); the deltoideus medialis (DEM); the deltoideus posterior (DEP); the trapezius superior (TRU); the rhomboid major (RHO); the brachioradialis (BRA); the brachialis (BRC); the supinator (SUP); the pronator teres (PTE); the pectoralis major (PEC); the infraspinatus (INF); the teres major (TEA).

sEMG was recorded from both ULs separately, while the patient was performing 7 voluntary unilateral UL motor tasks (10 repetitions each), for a total of 70 trials, in interaction with the VRRS® system (Khymeia Group Ltd., Padova, Italy). Each task involved a standardized trajectory that the patient was asked to emulate. The trajectories, first pre-recorded using the system and then displayed in a virtual environment (29), included: UL elevation, UL elevation with restriction, UL forward reaching, shoulder abduction, forearm pronation-supination, shoulder internal-rotation, shoulder external-rotation.

### sEMG feature extraction

We extracted the muscle synergy features from the sEMG signals following the pre-processing and extraction procedures described in previous studies (13, 20). We extracted muscle synergies from both the affected and unaffected ULs separately, by decomposing the signals chained across tasks using the Non-negative Matrix Factorization (NMF) algorithm. Before extraction, we pre-processed the sEMG signals through the following steps: band-pass filtering (10–500 Hz), normalization to unit variance, rectification and finally low-pass filtering at 12 Hz (30). We selected the number of muscle synergies using cross-validation for sEMG reconstruction factor, with a threshold of R^2^ > 80% (13).

After synergies extraction we calculated, for each participant, the following muscle synergies parameters: (i) number of muscle synergies from the affected (N-aff) and unaffected (N-un) UL; (ii) number of synergies preserved (N-sh), defined as the number of pairs of synergies between the affected and unaffected arms with a scalar product > 0.8 between them, i.e. indicating good similarity; (iii) median scalar product (M-sp) between the affected and unaffected UL, considered as an informative index of similarity between the two ULs with higher values indicating more similarity (29). Then, we estimated merging (Me) and fractionation (Fr) indexes of the muscle synergies in the affected UL with respect to the total number of synergies extracted from the unaffected UL, using the procedure from Cheung et al. (13). In this definition, merged synergies are affected-side synergies that can be described as linear combinations of two or more unaffected side synergies. The merging index is a nonnegative coefficient denoting the degree of contribution of a specific unaffected-arm synergy to the structure of a specific affected-arm synergy. Merging indexes were extracted using nonnegative least squares (function lsqnonneg in Matlab). As in (13), also in our case an unaffected-arm synergy was defined to contribute to the merging of an affected-arm synergy if its merging index was >0.2.

Similarly, fractionation is defined in the same way as merging after swapping the roles of affected-arm and unaffected-arm synergies. Affected-arm synergies presenting fractionation are those that, when linearly combined with one or more affected-arm synergy, constitute an unaffected arm synergy. Fractionation indexes were also extracted using nonnegative least squares. An unaffected-arm synergy was defined as a fractional part of an affected-arm synergy if its fractionation index was >0.2.

### Statistical analysis

The characteristics of the sample were reported by descriptive statistics (i.e., mean, standard deviation, median, interquartile range, absolute and percentage frequencies) for demographics, clinical and muscle synergies parameters. Moreover, the sample was stratified according to the recovery phase and the level of UL motor impairment at baseline. Respectively, the recovery phase after stroke was stratified according to the ESO guidelines as early subacute (from 7 days to 3 months), late subacute (from 3 to 6 months) and chronic (after 6 months) (7); while the level of UL motor function impairment according to Cheung et al. as severe (0 < FMA-UE < 31 points), or mild (FMA-UE ≥ 31 points) (13).

To determine whether the treatment induced relevant changes in outcome measures, we compared the score after treatment with baseline, considering as minimally clinically important difference (MCID) a change higher than 5 points at the FMA-UE (31). According to this criterion, participants were stratified as responding (Responder) or not (Non-Responder) to the treatment and differences between these subgroups were investigated both at the clinical and muscles synergy levels.

The Shapiro-Wilk Test was used to test data distribution properties for all the variables, then parametric (e.g., T-test) or non-parametric tests (e.g., Wilcoxon Test) were chosen accordingly. One-way analysis of variance (ANOVA) was used to test differences in the merging index according to treatments received, corrected post-hoc by the Tukey test for pairwise, to determine mean differences between pairs of treatments. Statistical significance level was set to 0.05 for all the tests.

### Synergies Clustering and Functions

To investigate whether specific muscle synergies correlated with patients’ characteristics, we derived a series of synergy “Vocabularies” from sub-datasets of muscle synergies relative to different conditions and patient subgroups. The vocabularies were constituted by synergy prototypes derived from a cluster analysis aiming at grouping together similar synergies from all the synergies in each sub-dataset. We identified three synergies vocabularies: the first vocabulary was derived from the synergies extracted from the affected UL before therapy (Vpre), the second vocabulary was derived from the synergies extracted from the affected UL after therapy (Vpost) and the third was derived from the synergies extracted from the unaffected UL at both timepoints (Vunaff). The clustering prototypes were derived by applying a hierarchical clustering algorithm based on Ward’s distance (32) to the complete synergies dataset relative to the sub-group analysed. The optimal number of clusters for each analysis was selected using the Silhouette algorithm (33).

After the derivation of the vocabularies, we characterized each vocabulary based on the UL motor functions it represented. The aim was to characterize whether the number of individual synergies associated with a certain motor function (e.g., elbow flexion) changed due to impairment and /or recovery. The synergies-to-function characterization assumed that each muscle in each specific synergy actively contributes its associated biomechanical function to the synergy’s motor function (e.g., biceps muscles are associated with elbow flexion, pectoralis major is associated with shoulder flexion and adduction). We identified 8 motor functions based on the degrees of freedom captured by the function-specific tasks adopted in the protocol and the muscles we recorded: shoulder flexion, shoulder extension, shoulder abduction, shoulder adduction, elbow flexion, elbow extension, forearm pronation, forearm supination.

One synergy was associated with a function if the average weights of all the muscles mapping to a specific function in that synergy was above a predefined threshold. The threshold was determined by first calculating the average synergy weight associated to all functions in all synergies. Then we calculated the maximum average weight of any function observed through all the synergies. The threshold for associating a function to a synergy was then set as the smaller maximum average weight of any function observed through all the synergies. In this way, each synergy is associated to at least one function, but most synergies map into more than one function. This characterization was done for all synergies vocabularies (Vpre, Vpost, Vunaff). After the synergy-to-function characterization we calculated the average number of functions associated to each synergy in each vocabulary. Finally, each single synergy extracted from the patients was associated with its corresponding synergy prototype in the vocabularies during the clustering analysis. This information was used to characterize the distribution of functions observed by the synergies sets of the different patient groups. Specifically, we evaluated the average number of functions per synergy for the Responders and Non-Responders subgroups for all three synergies vocabularies.

## Results

### Sample characteristics

We included 62 patients, all of them receiving all the treatment sessions in their respective group. With regard to the distance from stroke onset (7), 30 patients were in the early subacute phase, 9 patients in the late subacute phase, and 23 patients in the chronic phase. Finally, considering the FMA-UE MCID score (31), 34 were classified ad Responder and 28 as Non-Responder. The full patients’ characteristics are reported in Table 1.

**Table 1.** Demographic and clinical characteristics of the patients.

| **Patients (N = 62)** |  |
| --- | --- |
| Sex, males / females | 42 (68%) / 20 (32%) |
| Age, years | 62.02 (±13.68) |
| Diagnosis, ischemic / haemorrhagic | 50 (81%) / 12 (19%) |
| Hemisphere, left / right | 30 (48%) / 32 (52%) |
| Time-stroke, months | 13.95 (± 30.54) |
| Recovery phase, early subacute / late subacute / chronic | 30 (48%) / 9 (15%) / 23 (37%) |
| Level of impairment, severe / mild | 18 (29%) / 44 (71%) |
| Type of treatment, CT / VRT / RT | 8 (13%) / 34 (55%) / 20 (32%) |
| Treatment output, responder / non-responder | 34 (55%) / 28 (45%) |

*Values are expressed as mean ± standard deviation (SD) for quantitative measures, and number (n) and frequency percentages (%) for all the discrete variables. Abbreviation= CT: Conventional Treatment; VRT: Virtual Reality Treatment; RT= Robotic-assisted Treatment.*

### Clinical outcome

Overall, all the clinical outcome measures significantly changed after treatment, with the exclusion of the FMA-P. On average, the FMA-UE increased significantly by 7.14 (± 7.46, p< 0.001) points and considering the FMA-UE MCID (i.e., > 5 points) (31), the group of pooled participants had a clinically meaningful change of the upper UL function after treatment (Table2).

**Table 2.** Clinical outcome measures at baseline and after treatment.

| Clinical outcomes measures | T0<br>(n = 62) | T1<br>(n = 62) | $\Delta$<br>(T1 – T0) | P-value |
| --- | --- | --- | --- | --- |
| FMA-UE | 39.82 ± 15.22<br>41.5 [24.5] | 46.97 ± 13.41<br>48 [21] | 7.14 ± 7.46<br>6.5 [8] | < 0.001* |
| FMA-S | 20.35 ± 4.58<br>22 [5] | 22.02 ± 3.24<br>24 [3.75] | 1.66 ± 3.52<br>0 [3] | < 0.001* |
| FMA-P | 43.61 ± 4.31<br>45 [6] | 44.44 ± 4.01<br>45 [5.75] | 0.82 ± 3.50<br>0 [3] | 0.061 |
| MAS | 3.45 ± 3.61<br>2 [5.75] | 2.53 ± 3.21<br>1.5 [3] | -0.92 ± 2.46<br>0 [1] | 0.002* |
| FIM | 98.44 ± 18.88<br>102 [24.25] | 108.1 ± 17.77<br>114 [19] | 9.67 ± 10.76<br>8 [10] | < 0.001* |

Values are expressed as Mean ± SD and median [IQR]. FMA-UE= Fugl-Meyer Assessment- Upper Extremity; FMA-S= Fugl-Meyer Assessment-sensitivity; FMA-P= Fugl-Meyer Assessment- Pain and ROM; MAS= Modified Ashworth Scale; FIM= Functional Independence Measure; “ $\Delta$ ” = change achieved after treatment. P values < 0.05 using Wilcoxon Test. (\*) = statistically significance.

We compared the clinical results between Responders and Non-Responders patients (Table3).

**Table 3.**
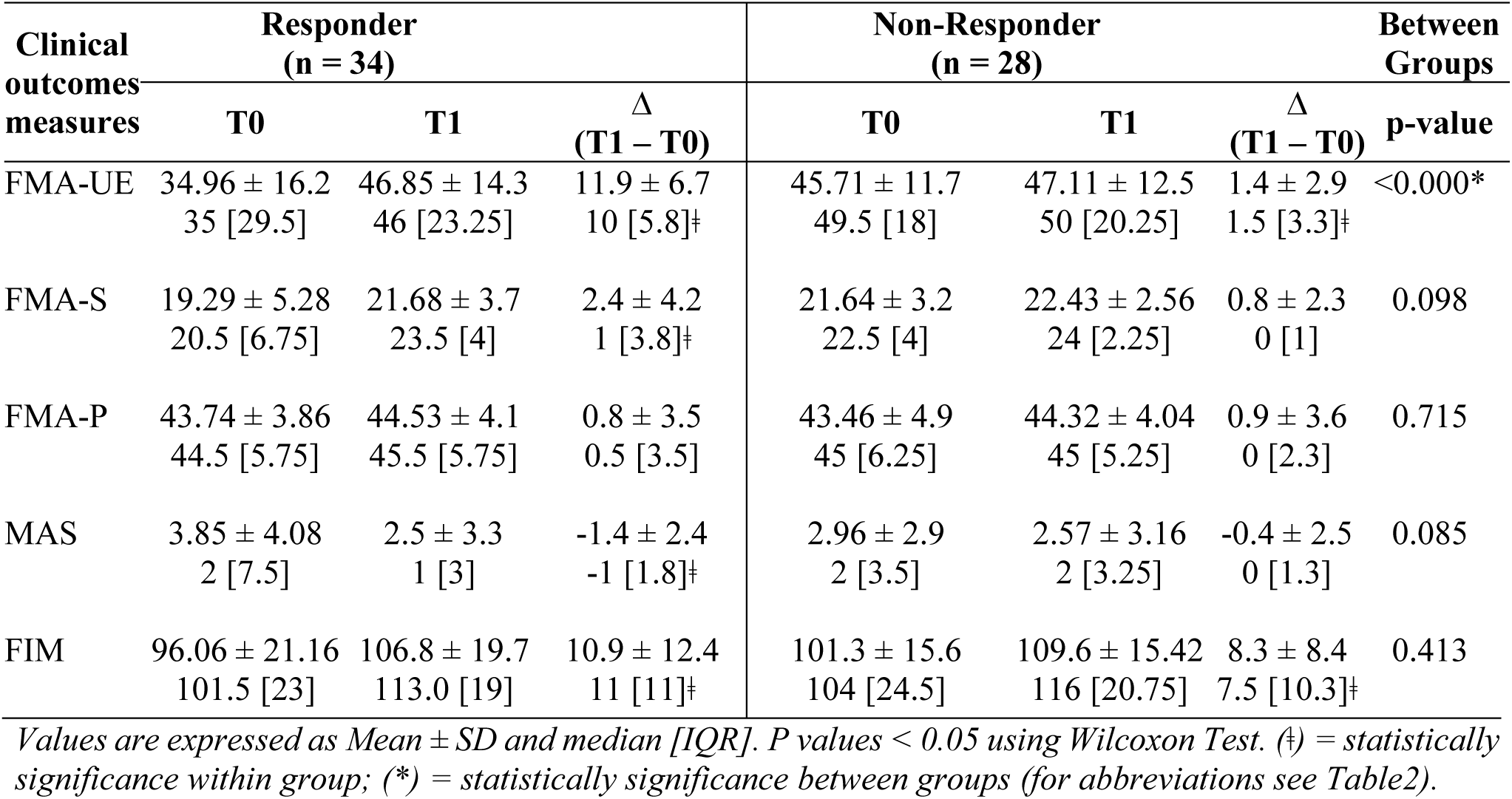
Clinical outcome measures at baseline and after treatment in Responder and Non-Responder subgroups.

| Clinical outcomes measures | Responder<br>(n = 34) |  |  | Non-Responder<br>(n = 28) |  |  | Between Groups<br>p-value |
| --- | --- | --- | --- | --- | --- | --- | --- |
| | T0 | T1 | $\Delta$<br>(T1 – T0) | T0 | T1 | $\Delta$<br>(T1 – T0) | |
| FMA-UE | 34.96 ± 16.2<br>35 [29.5] | 46.85 ± 14.3<br>46 [23.25] | 11.9 ± 6.7<br>10 [5.8]† | 45.71 ± 11.7<br>49.5 [18] | 47.11 ± 12.5<br>50 [20.25] | 1.4 ± 2.9<br>1.5 [3.3]† | <0.000* |
| FMA-S | 19.29 ± 5.28<br>20.5 [6.75] | 21.68 ± 3.7<br>23.5 [4] | 2.4 ± 4.2<br>1 [3.8]† | 21.64 ± 3.2<br>22.5 [4] | 22.43 ± 2.56<br>24 [2.25] | 0.8 ± 2.3<br>0 [1] | 0.098 |
| FMA-P | 43.74 ± 3.86<br>44.5 [5.75] | 44.53 ± 4.1<br>45.5 [5.75] | 0.8 ± 3.5<br>0.5 [3.5] | 43.46 ± 4.9<br>45 [6.25] | 44.32 ± 4.04<br>45 [5.25] | 0.9 ± 3.6<br>0 [2.3] | 0.715 |
| MAS | 3.85 ± 4.08<br>2 [7.5] | 2.5 ± 3.3<br>1 [3] | -1.4 ± 2.4<br>-1 [1.8]† | 2.96 ± 2.9<br>2 [3.5] | 2.57 ± 3.16<br>2 [3.25] | -0.4 ± 2.5<br>0 [1.3] | 0.085 |
| FIM | 96.06 ± 21.16<br>101.5 [23] | 106.8 ± 19.7<br>113.0 [19] | 10.9 ± 12.4<br>11 [11]† | 101.3 ± 15.6<br>104 [24.5] | 109.6 ± 15.42<br>116 [20.75] | 8.3 ± 8.4<br>7.5 [10.3]† | 0.413 |

Values are expressed as Mean ± SD and median [IQR]. P values < 0.05 using Wilcoxon Test. (†) = statistically significance within group; (\*) = statistically significance between groups (for abbreviations see Table2).

The scatter plot in Figure 1 shows the relationship between the amount of UL motor recovery (i.e., ΔFMA-UE) and the baseline level of UL motor impairment (i.e., FMA-UE baseline score). Patients in the Responder subgroup expressed higher recovery for low baseline scores, with the highest FMA-UE improvement for patients with severe motor impairment (i.e., FMA-UE < 20 points). Conversely, patients in the Non-Responder group had a similar change-score regardless the baseline score (Figure 1).

**Figure 1.**
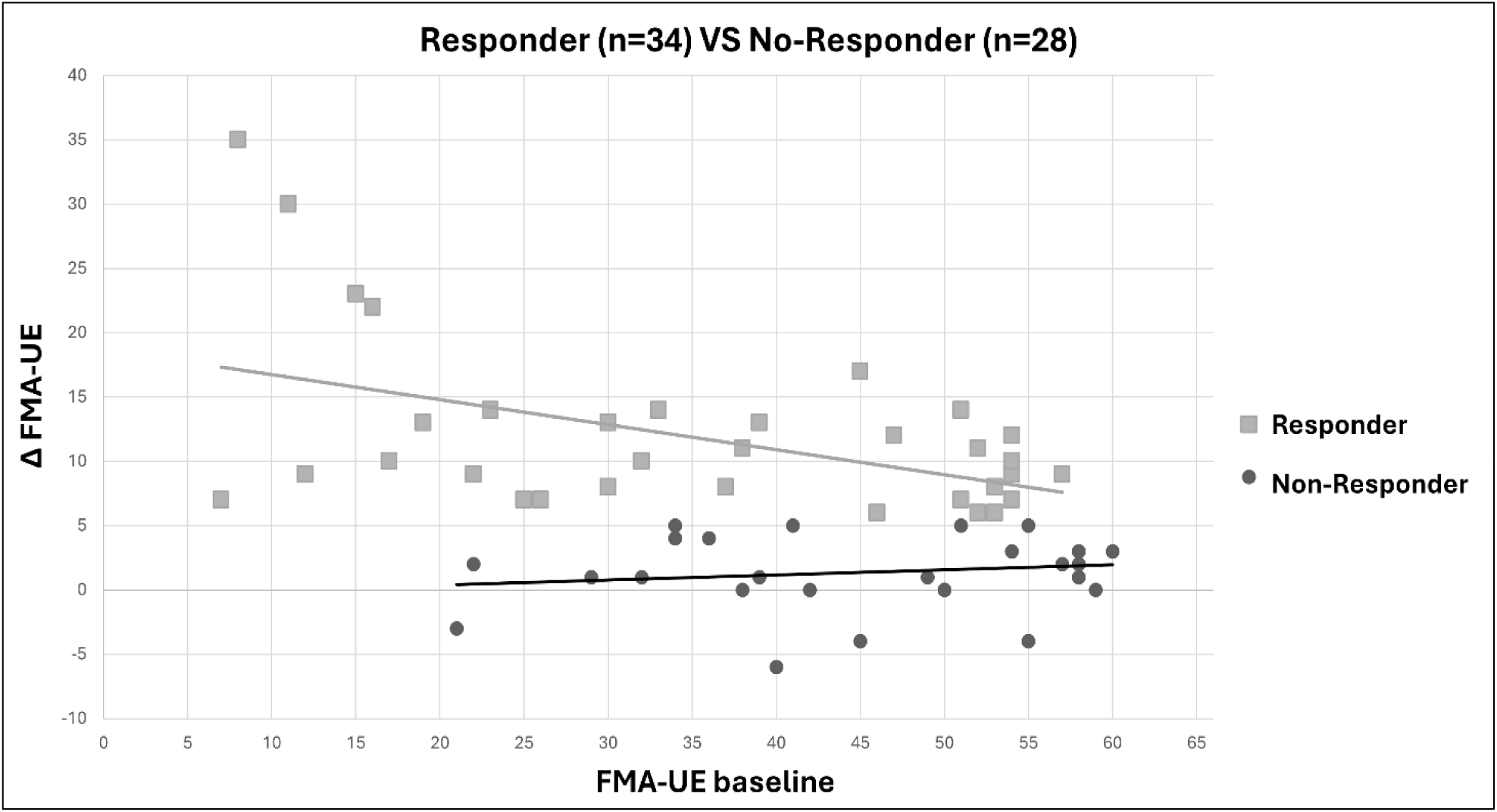
Relationship between the motor impairment at baseline and the amount of recovery in Responder and Non-Responder subgroups. Each symbol represents a participant, from Responder subgroup (squares) and Non-Responder subgroup (dots). On the x-axis, the Fugl-Meyer Upper Extremity score at baseline and on the y-axis the delta (Δ) between the final score and the baseline. Across all patients (No.=62) we found a significant negative correlation (Spearman‘s rank correlation, rho=-0.373, *95%CI [−0.136, −0.569],* p-value = 0.0029). Thus, we separated the results in Responder (Spearman‘s rank correlation, rho=-0.325, *95%CI [0.15, −0.598],* p-value = 0.0606) and Non-Responders (Spearman‘s rank correlation, rho=0.128, *95%CI [0.478, −0.258],* p-value= 0.5175).

### Muscle synergies parameters

We analysed the change of muscle synergies parameters for the patients as a whole group and in the subgroups of Responders and Non-Responders separately. Overall, the number of modules was comparable between the affected and unaffected UL at baseline (p = 0.107).

After treatment no difference in N-aff was retrieved (p = 0.374), while M-sp improved significantly (p = 0.004), suggesting that treatment makes the synergies of the impaired UL more similar to those of the unimpaired UL. We did not observe significant changes in Me(%) and Fr(%) indices after treatment (Table 4).

**Table 4.** Muscle synergies parameters results, before and after the treatment for all patients (n=62).

| Muscle Synergies parameters | T0<br>(n = 62) | T1<br>(n = 62) | $\Delta$<br>(T1 – T0) | p-value |
| --- | --- | --- | --- | --- |
| N-aff | 8.82 ± 1.49<br>9 [2] | 8.0 ± 1.37<br>9 [1] | -0.16 ± 1.15<br>0 [2] | 0.374 |
| N-un | 8.61 ± 1.28<br>9 [1.75] | 8.66 ± 1.04<br>9 [1] | 0.05 ± 1.05<br>0 [2] | 0.716 |
| N-sh | 6.77 ± 1.43<br>7 [2] | 6.73 ± 1.46<br>7 [2] | 0.00 ± 1.52<br>0 [2] | 0.885 |
| M-sp | 0.88 ± 0.07<br>0.88 [0.11] | 0.90 ± 0.06<br>0.92 [0.08] | 0.03 ± 0.06<br>0.02 [0.07] | 0.004* |
| Me (%) | 59.26 ± 20.22<br>58.55 [25.6] | 53.40 ± 20.81<br>50 [26.67] | -5.86 ± 32.56<br>-7.02 [26.67] | 0.068 |
| Fr (%) | 7.34 ± 10.4<br>0 [11.11] | 7.27 ± 9.34<br>0 [11.11] | -0.07 ± 11.49<br>0 [17.67] | 0.952 |

Values are expressed as Mean ± SD and median [IQR]. N-aff= number of synergies extracted from affected upper limb; N-un= number of synergies extracted from unaffected upper limb; N-sh= number of synergies shared between upper limbs; M-sp= median of scalar product; Me= merging index; Fr= fractionation index; “ $\Delta$ ” = change achieved after treatment. P values < 0.05 using Wilcoxon Test. (\*) = statistically significance.

The merging index at the baseline (T0) showed a similar correlation for the Responder (y = −0.477 x + 80.,04; R² = 0.133) and for Non-Responder groups (y = −0.028x – 54.88; R² = 0.025) (Figure 2) as the previous plot (Figure 1). This suggested that Me(%) index might represent a surrogate neurophysiological feature based on muscle synergies describing clinical functional recovery.

**Figure 2.**
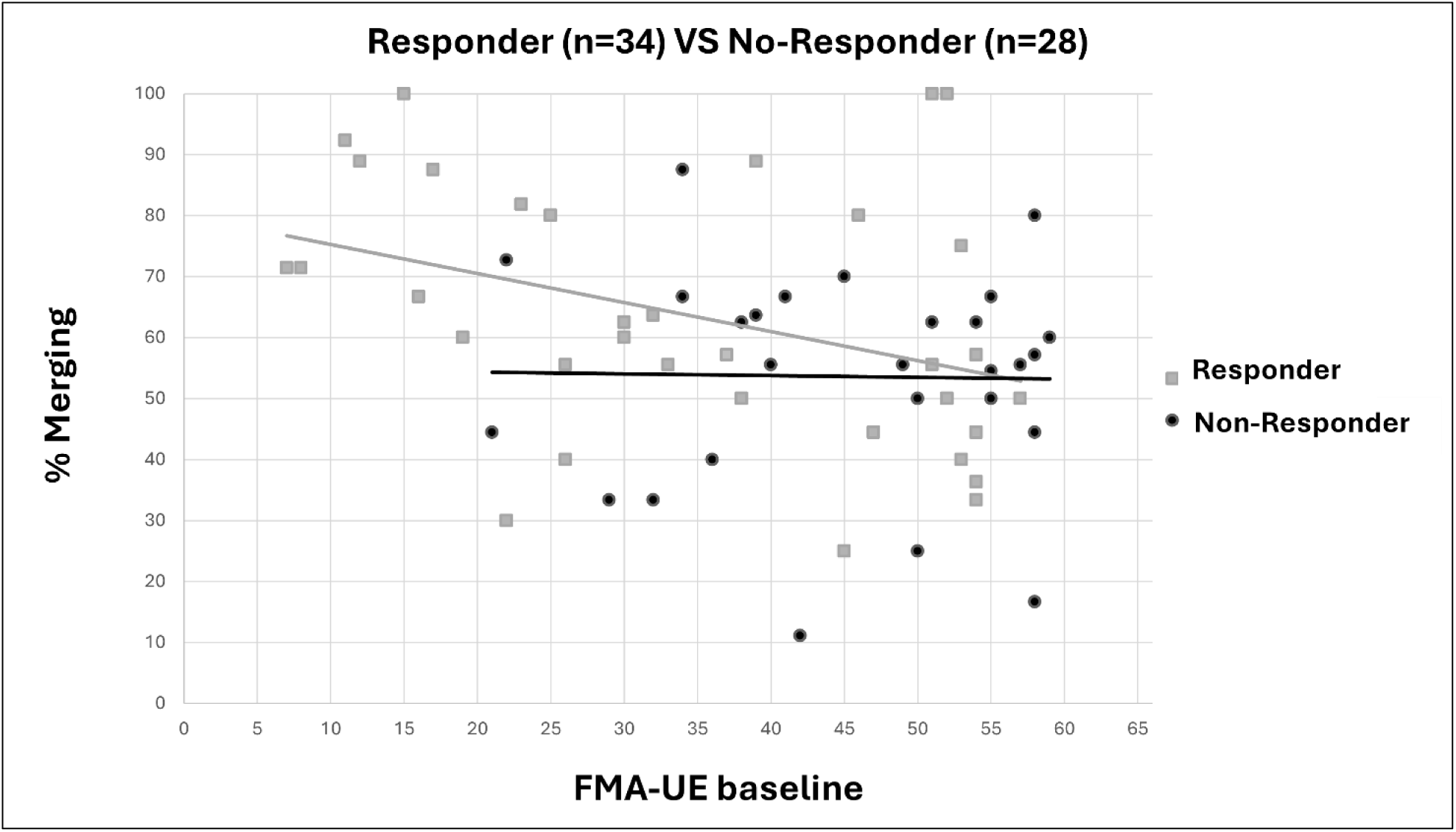
Relationship between the motor impairment at the baseline and the amount of merging at the baseline in Responder and Non-Responder subgroups. Each symbol represents the FMA-UE and % of merging at baseline for a single participant belonging the Responder (squares) or Non-Responder (dots) subgroup. Across all patients (No.=62) we found a significant negative correlation (Spearman‘s rank correlation, rho=-0.275, *95%CI [−0.028, −0.491],* p-value = 0.030). Thus, we separated the results in Responders (Spearman‘s rank correlation, rho= −0.438, *95%CI [−0.118, −0.676]*, p-value = 0.009) and Non-Responders (Spearman‘s rank correlation, rho=0.042, *95%CI [0.409, −0.336]*, p-value= 0.832).

At the level of Responder and Non-Responder subgroups, N-aff (p = 0.552), Me(%) (p = 0.171) and Fr(%) (p = 0.136) were comparable between subgroups, at baseline, with only Me (%) changing between groups significantly (p = 0.025), indicating that impaired synergy modules in the Responder subgroup were no longer described as merged modules of the unaffected UL (Table 5).

**Table 5.** Change of muscle synergies after treatment, in the Responders and Non-Responder subgroups.

| Muscle Synergies parameters | Responder (n = 34) |  | Non-Responder (n = 28) |  | Between Groups p-value |
| --- | --- | --- | --- | --- | --- |
|  | T0 | T1 | T0 | T1 |  |
| N-aff | 8.94 ± 1.54<br>9 [2] | 8.59 ± 1.31<br>9 [1] | 8.68 ± 1.44<br>9 [1.25] | 8.75 ± 1.46<br>9 [2] | 0.180 |
| N-un | 8.71 ± 1.36<br>9 [2] | 8.68 ± 1.17<br>9 [1] | 8.5 ± 1.2<br>8.5 [1] | 8.64 ± 0.87<br>9 [1] | 0.586 |
| N-sh | 6.94 ± 1.30<br>7 [2] | 6.79 ± 1.49<br>6.5 [2] | 6.54 ± 1.58<br>7 [1] | 6.65 ± 1.44<br>7 [1.75] | 0.269 |
| M-sp | 0.87 ± 0.07<br>0.89 [0.12] | 0.90 ± 0.06<br>0.92 [0.1] | 0.88 ± 0.06<br>0.89 [0.08] | 0.91 ± 0.06<br>0.92 [0.09] * | 0.181 |
| Me (%) | 63.37 ± 21.15<br>60 [30] | 51.17 ± 22<br>50 [29.2] * | 54.27 ± 18.1<br>56.4 [22.3] | 56.11 ± 19.3<br>52.27 [30.2] | 0.025* |
| Fr (%) | 5.67 ± 8.49<br>0 [10] | 7.67 ± 8.58<br>4.5 [13.8] | 9.35 ± 12.2<br>10.1 [12.5] | 6.80 ± 10.34<br>7 [11.11] | 0.070 |

Values are expressed as Mean ± SD and median [IQR]. N-aff= number of synergies extracted from affected upper limb; N-un= number of synergies extracted from unaffected upper limb; N-sh= number of synergies shared between upper limbs; M-sp= median of scalar product; Me= merging index; Fr= fractionation index. $P$ values < 0.05 using Wilcoxon Test. (\*) = statistically significance.

Considering each subgroup, results showed that percentage of merging (Me%) decreased significantly from T0 to T1 in the Responder subgroup (p = 0.016), whereas the similarity index (M-sp) significantly increased in the Non-Responder subgroup (p = 0.006).

In addition, we compared the change of merging with regard to the different training modalities. We carried out a one-way ANOVA test showing a significant effect of training modalities on changes of Me(%) (F = 3.52; p = 0.036), specifically confirmed for VRT and RT treatments after post-hoc Tukey correction (F = 18.8 (1.4-36.1), p = 0.031), although the baseline Me(%) value were significantly different between the VRT and RT groups (i.e., VRT = 52.59%; RT = 69.66%; p = 0.001).

We identified three sets of synergy vocabularies from the synergies extracted, across all patients, from the affected arm before (Vpre) and after (Vpost) therapy, and from the unaffected arm at both timepoints (Vunaff), respectively. We found that Vpre was constituted by 13 synergies (Figure 3), Vpost by 14 synergies (Figure 4), while the Vunaff was constituted by 16 synergies (Figure 5).

**Figure 3.**
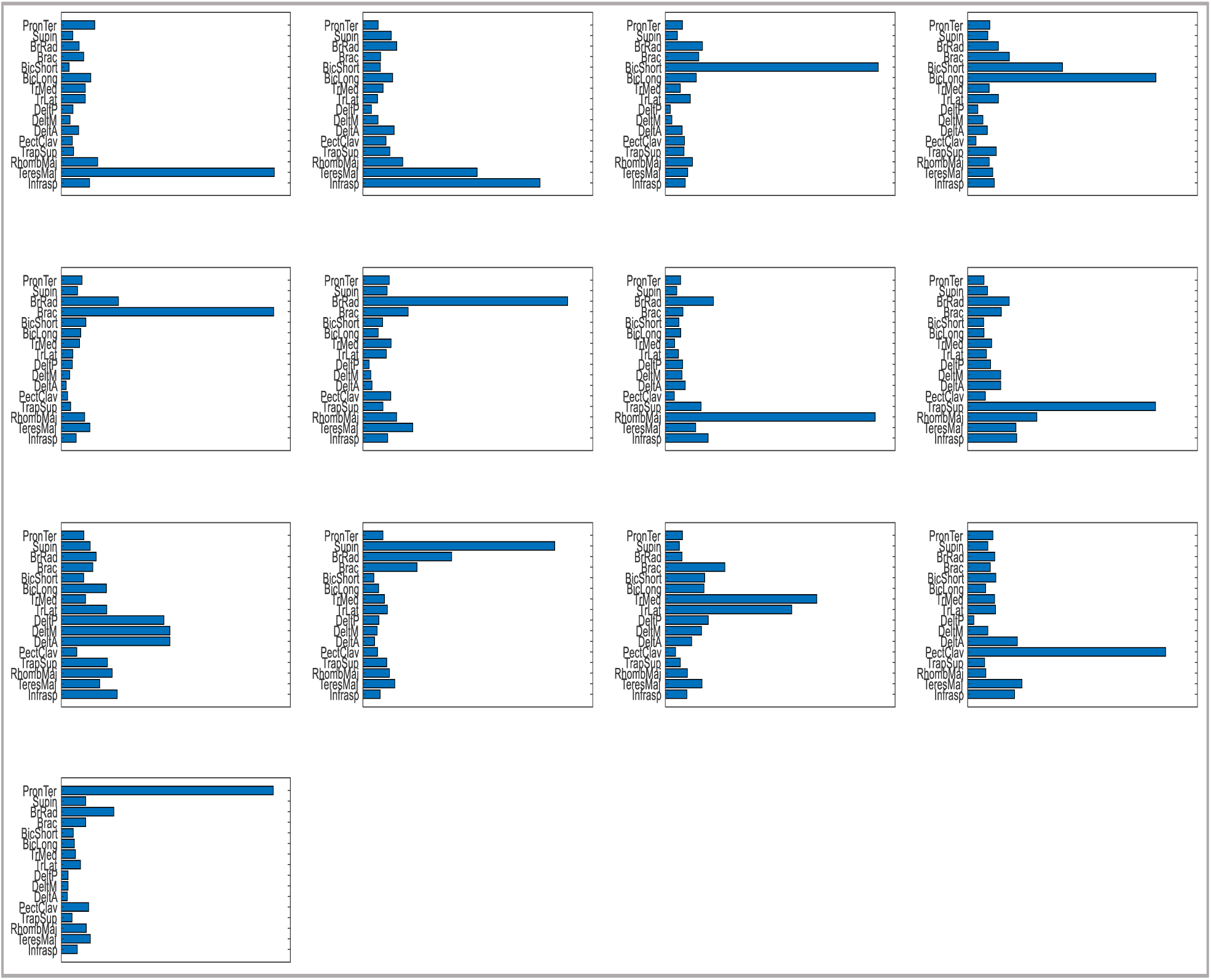
Vocabularies of muscle synergies in affected limb at the baseline (T0). The representation of each muscle synergy; from the top of the plot, the muscle are the following: PronTer= pronator teres; Supin= supinator; BrRad= brachioradialis; Brac= brachialis; BicShort= biceps brachii, short; BicLong= biceps brachii, long; TrMed= triceps brachii medial; TrLat= triceps brachii lateral; DeltP= deltoideus posterior; DeltM= deltoideus medialis; DeltA= deltoideus anterior; PectClav= pectoralis major; TrapSup= trapezius superior; RhombMa= rhomboid major; TeresMA= teres major; Infrasp= infraspinatus.

**Figure 4.**
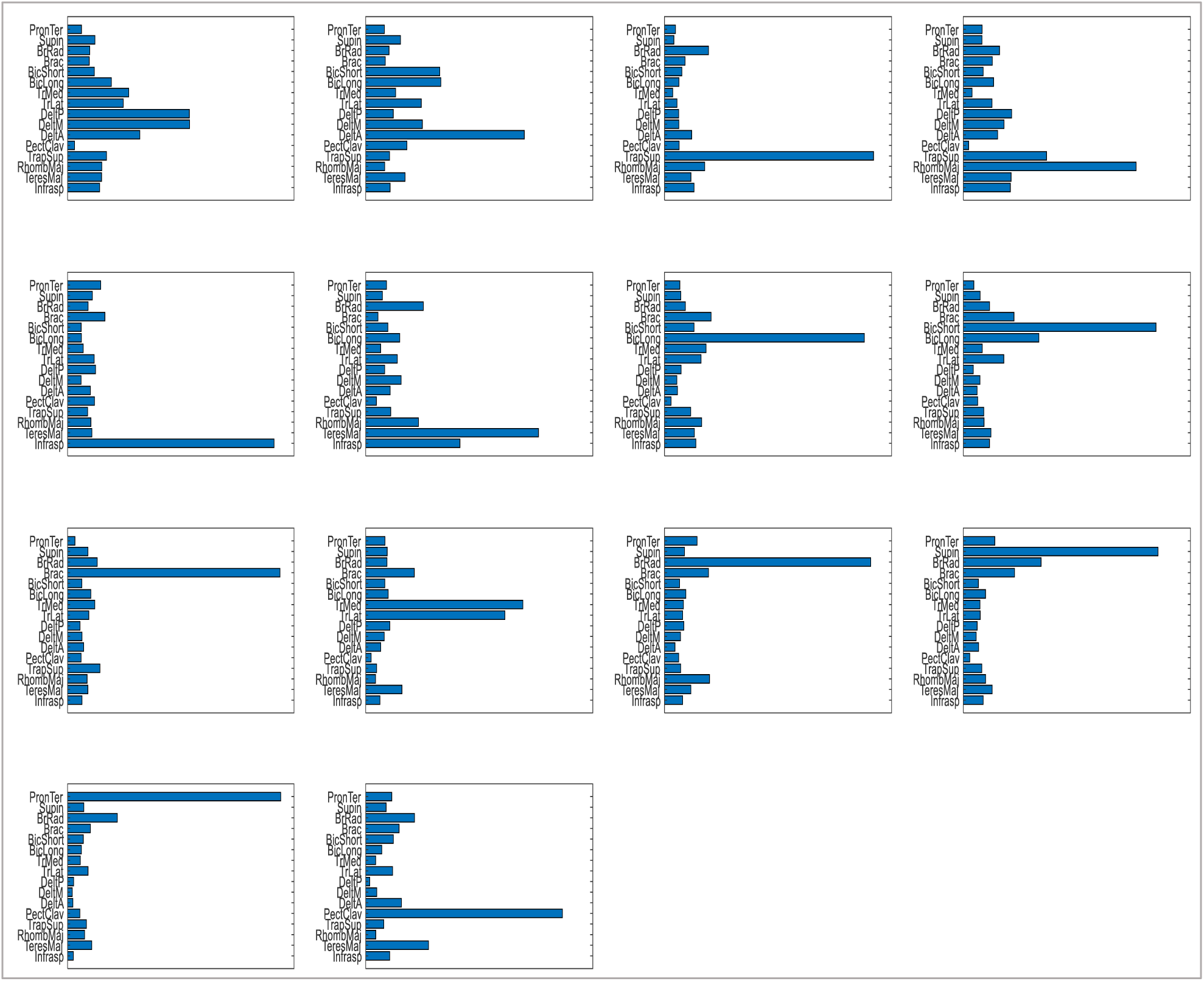
Vocabularies of muscle synergies in affected limb after treatment (T1). The representation of each muscle synergy (for abbreviations see Figure3).

**Figure 5.**
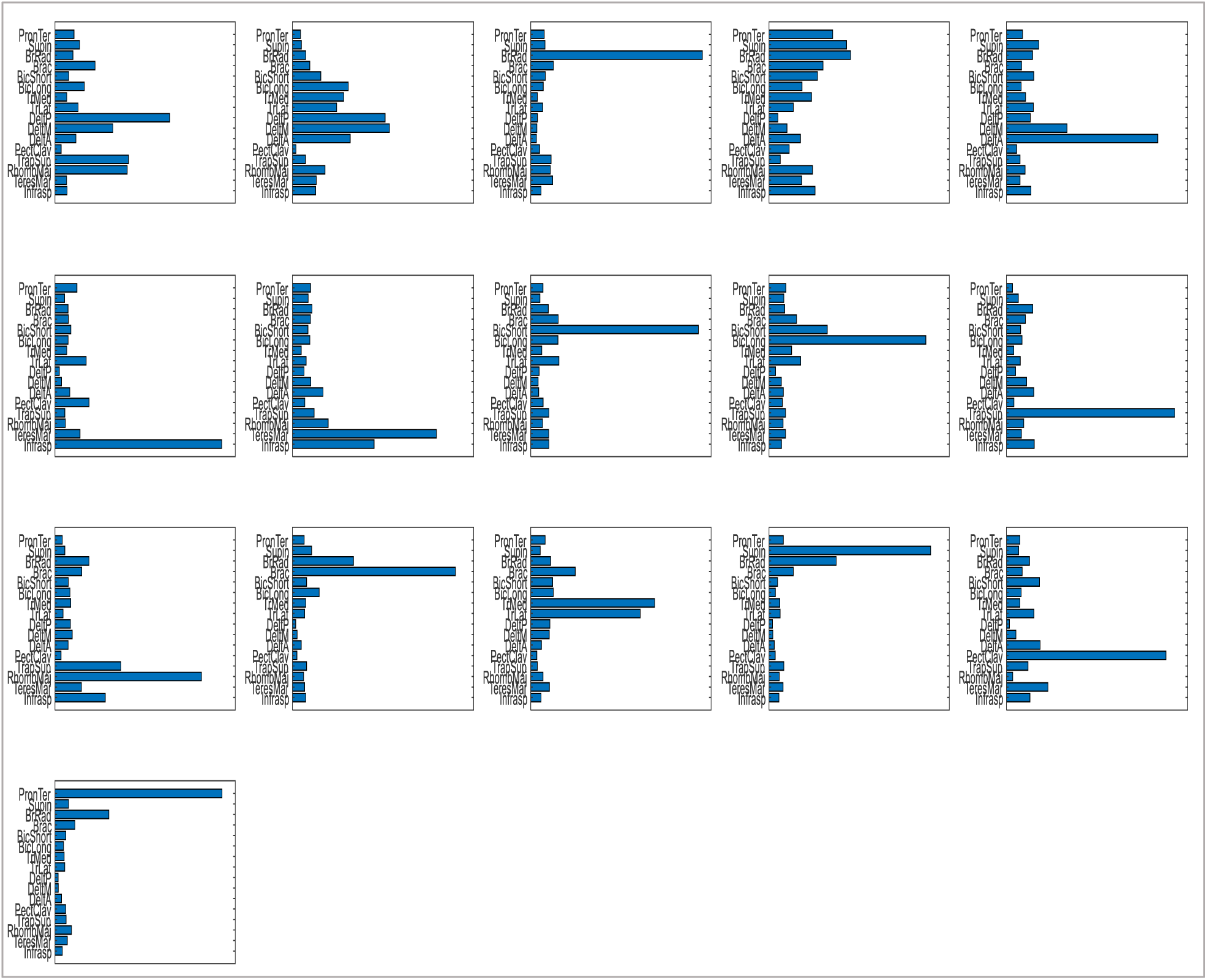
Vocabularies of muscle synergies in the unaffected limb, before and after the treatment (T0 and T1). The representation of each muscle synergy (for abbreviations see Figure3).

We associated one or more UL motor functions to each synergy that composed the vocabularies, and we analysed how many functions on average were associated with each synergy.

Results showed that Vunaff synergies were associated with an average of 1.33 functions per synergy across all participants.

On average, Vpre synergies were associated with 2.54 and 2.61 functions, while Vpost synergies with 1.37 and 1.42 functions in the Responder and No-Responder subgroups, respectively. Finally, we analysed how the UL motor functions used were expressed across all the synergies, before and after therapy, in the entire sample and in both subgroups of Responder and Non-Responder, comparing it with the unaffected arms (Figure 6).

**Figure 6.**
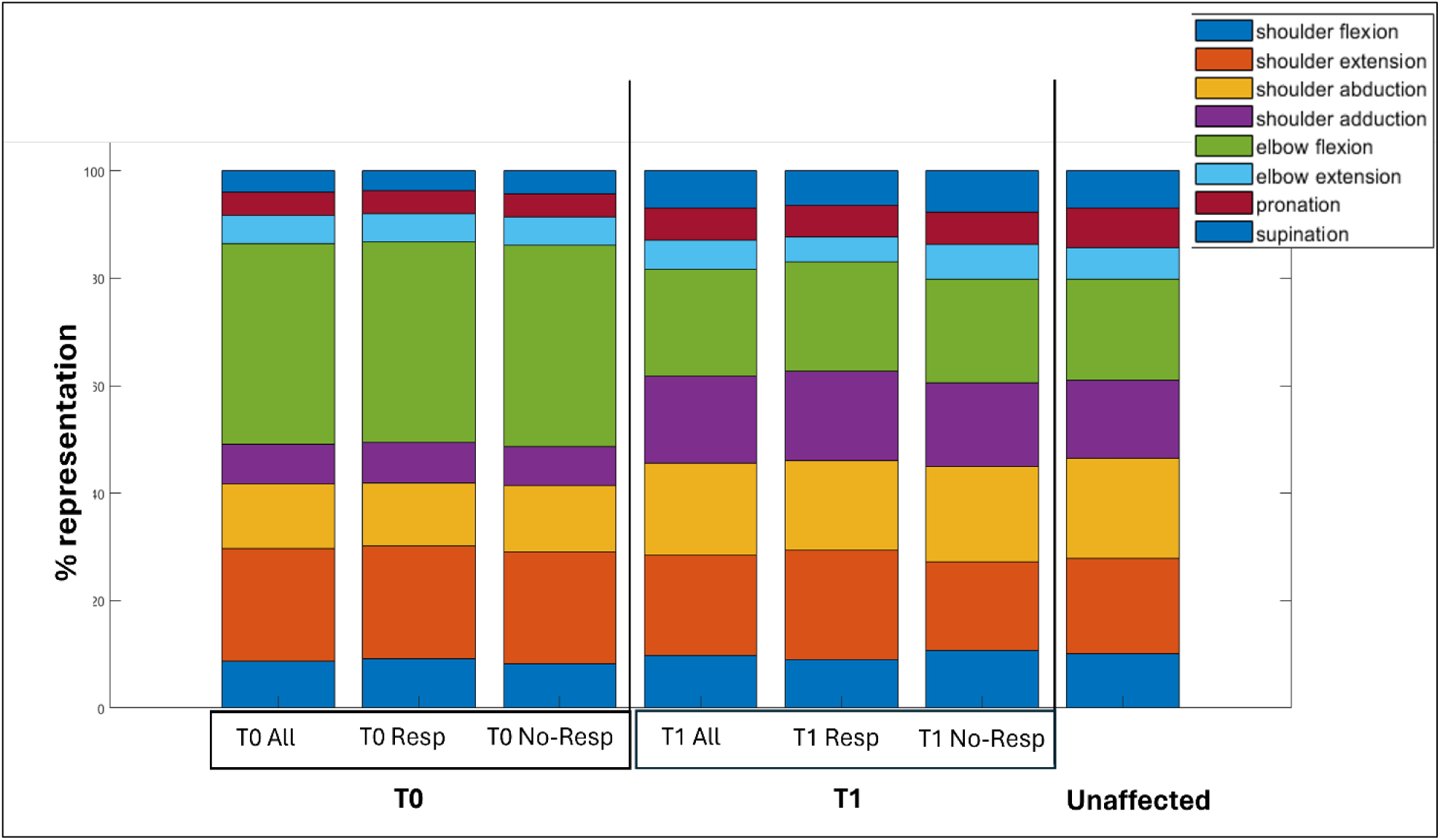
The function representation map of UL functions, in three Vocabularies. The representation of each Vocabulary of muscle synergies of affected upper limb for all patients (All), in Responder (Resp) and Non-Responder (No-Resp) subgroup, before (T0, Vpre) and after (T1, Vpost) treatment; then, the Vocabulary of muscle synergies of unaffected upper limb (Unaffected).

Results showed no substantial differences between the functions of Responder and Non-Responder subgroups, before the therapy. After therapy, shoulder extension (T1 Responders= 20.46%; T1 No-Responders= 16.51%) and adduction (T1 Responders= 16.62%; T1 No-Responders= 17.74%) were slightly more represented in the Responder subgroup, while shoulder flexion (T1 Responders= 8.95%; T1 No-Responders= 10.70%) and elbow extension (T1 Responders= 4.60%; T1 No-Responders= 6.42%) were more represented in Non-Responder subgroup.

Generally, the comparison between affected and unaffected ULs showed that after therapy the functional representation of impaired UL was more similar to those of the unimpaired UL, conveying the same information as the merging index analysis.

## Discussion

In this study we compared clinical outcomes and muscle synergies parameters in the characterization of motor recovery after specific UL motor rehabilitation in stroke survivors. We extracted synergy-related parameters, including merging and fractionation indices (13), to investigate different levels of motor impairment and different amount of motor recovery distinguishing between patients who responded to rehabilitation and those who did not, based on MCID thresholds of the FMA-UE (31).

Our results showed a significant difference in the merging index between Responders and Non-Responder subgroups, with Responders showing a higher merging before rehabilitation and a significant decrease of merging after therapy (p = 0.016). This finding confirmed findings from Pan et al. that investigated the relationship between merging coefficient and level of UL motor impairment in a population of stroke survivors, describing the merging process as the mechanism explaining the characteristics of disrupted muscle synergies (34, 35). These results disclosed the role of rehabilitation in promoting a “de-merge” effect on muscle synergies of the affected limb, in patients expressing functional recovery at clinical outcome measures, after stroke. Thus, the merging index at baseline could be considered a potential prognostic biomarker for characterising motor recovery, allowing the identification of patients that are more likely to respond to rehabilitation interventions (36).

In our study, the fractionation index was not informative, even when considering different time after stroke (i.e., before and after 6 months). Cheung et al. found that the fractionation index was higher in chronic patients, possibly indicating that a longer time of rehabilitation practice might induce new motor strategies supporting recovery of autonomy in activities of daily living (13). In our sample, the time from lesion onset was heterogenous, with higher percentage of patients in the early and late-subacute phase. In future research, we should investigate fractionation index in a wider sample of patients also including subjects in the chronic phase of recovery (i.e., after 6 months from stroke onset). Moreover, we may investigate other groups of muscles (e.g., trunk dorsal muscles) and their contribution during execution of UL tasks. In such analyses, the fractionation index could be a more informative feature for identification of changes in postural control or motor compensation strategies (37, 38).

Considering the whole sample, functional improvement was associated with substantial preservation of the number of muscle synergies and increased similarity between affected and unaffected ULs after treatment. Recently, Seo et al. confirmed that the number of synergies extracted did not change after a robotic-based training (17, 39). Nonetheless, only in patients responding to rehabilitation the muscle synergies composition changed, increasing the component of each muscle within synergies after the training (17). Moreover, in our previous results, we compared clinical, kinematic and electromyographic features extracted before and after UL robotic-based training and usual care (15, 40) in a sample of stroke survivors. Results showed that all the patients preserved organization of modules, but their structures, as described by similarity index between muscle synergies weightings or merging and fractionation indexes, were different following rehabilitation (13, 41). In this work similarly, we did not observe differences in the number of modules of the affected UL between patients responding or not to rehabilitation. Our results supported the evidence that patients expressed different amount of recovery (42). Further, to explore the potential in discriminating different muscle profiles for patients responding or not to UL rehabilitation, we employed an innovative approach based on cluster analysis and mapping the synergy clusters to their functional representations (Figure 6), defining muscle synergies Vocabularies for all the different scenarios (i.e., impaired UL before rehabilitation, impaired UL after rehabilitation, unaffected UL).

At baseline, motor functions in the affected UL did not characterize the Responder patients’ muscles profile. Indeed, all patients expressed an over-representation of the elbow flexion function and an under-representation of the shoulder adduction/abduction and pronation/supination functions at baseline, when compared with the not-affected UL model. The UL impaired muscle-synergies-based profiles confirmed results from other trials. For instance, the main differences in reaching tasks between severely impaired patients and healthy people, was the increased activation of pectoralis major muscle and elbow flexors (i.e., biceps brachii and brachioradialis), and the decreased activation of elbow extensors muscle (i.e., triceps brachii) (9, 34).

On the other hand, after therapy, the affected UL profile of muscle-synergies in both Responder and Non-Responder subgroups, became very similar to the unaffected UL profile, with potential small differences between subgroups. These results suggested that, regardless of whether patients respond or not to therapy, the modules reflect a functional re-organization that makes them more similar to their unaffected counterpart.

For both subgroups, after rehabilitation, the number of functions related to each synergy decreased. Thus, muscle synergies were more specialized because more synergies carried one function, and each synergy was more represented by one function. In particular, the elbow flexor muscles were less represented, while the activation of the back and shoulder muscles were more consistent. In summary, the cluster analysis did not show substantial differences between Responders and Non-Responders, but indicated an effect of rehabilitation towards “normalization” of UL muscles activation.

This study has several limitations, mainly due to the critical aspects of procedures adopted for muscle synergies extraction. Indeed, the number of muscle synergies depends on the acquisition protocol and on technical parameters used for features extraction (21, 43), highlighting the urgency to develop guidelines based on experts’ consensus (38, 43). Moreover, we defined a priori the threshold for classifying a clinically important improvement after rehabilitation treatment, based on the MCID of the FMA-UE from literature (31). This threshold might be different according to different individual characteristics, not considered in our sample. Future clinical trials should better identify the phenotype of people who benefit from specific motor rehabilitation interventions (4). Finally, the composition of Vocabularies has been defined based on not-affected biomechanical functions, which may be not the correct reference to be considered in impaired people, due to natural disease related alterations (44).

## Conclusions

In our study, we investigated whether muscle synergies parameters can describe motor recovery as defined by clinical outcome measures, in patients undergoing specific UL rehabilitation treatment after stroke. Results showed that patients expressing better motor recovery after rehabilitation showed a significant decreasing of merging, that became similar to the one of patients not responding to rehabilitation. These findings suggest that the level of merging may have potential for prognosis of UL motor recovery in patients taking advantage from the rehabilitation.

Finally, we described the synergies vocabularies of the affected and unaffected UL after stroke, showing that the functional profile of the affected UL became more similar to the unaffected limb after rehabilitation. These findings support the idea that a healthy model should be considered as a reference for the development of motor rehabilitation treatments, including those based on technology (45). Muscle synergies cluster analysis may support the clinical assessment to identify the pair of muscles to be trained or untrained, with the aim to personalize the rehabilitation program, fostering “restitution” or “compensation” of UL motor functions (46, 47). Further, the simultaneous collection of other instrumental data (e.g., kinematics) and the application of more advanced analysis, such as new algorithms, may be considered to provide more accurate insights in this field (48).

Finally, the muscle synergies-based approach may be considered as a reference standard model for the development of new rehabilitation technologies, considering that development of devices and motor control principles have computational commonalities to be solved when manufacturing (49). Therefore, the application of muscle synergies-based approach to technology development may be a strategy to provide intuitive and valid feedback to patients interacting with the device, with the aim to promote de-coupling of improperly co-activated muscles and to stimulate motor recovery based on good redundancy (39).

## Data Availability

All data produced in the present work are contained in the manuscript.

## Ethics approval and consent to participate

The study was conducted according to the guidelines of the Declaration of Helsinki, and approved by the Ethics Committee for Clinical Experimentation (CESC) of Venice and San Camillo IRCCS hospital (Prot. No. 2015.14) and the written informed consent from all participants enrolled.

## Authors’ contributions

GP, GS, AT contributed to the experimental process, manuscript drafting and reviewing. AT, GP, TL, MF and JJ contributed to the clinical trial design and the participant management. IC implemented the robot-training protocol in Milan. GP, DR, IC and TL participated in acquisition and processing the instrumented data. GP, LM and GS contributed to data analysis. MF and JJ coordinated the team in Milan. IC, TL, MF, JJ and VCKC critically reviewed the manuscript. AT was the PI of the grants No. GR-2011-02348942 and No. RF-2019-12371486 sustaining the trial. AT conceived the study and coordinated the whole projects. All Authors read and approved the final manuscript.

## Acknowledgements

We would like to acknowledge Marialuisa Bullo, Riccardo Spezzamonte, Michela Agostini, Alberto Marzegan, Thomas Bowman, Rita Bertoni and all physiotherapists and patients for their contribution to data collection.

